# What were the challenges and needs before implementing routine pulse oximetry in IMCI consultations at primary health centres in West Africa? Baseline site assessment of the operational AIRE project, 2020

**DOI:** 10.1101/2024.10.14.24315436

**Authors:** Gildas Boris Hedible, Gildas Anago, Severin Lenaud, Désiré Néboua, Zineb Zair, Abdoul Guaniyi Sawadogo, Sarah Louart, Valérie Zombré, Dieney Fadima Kaba, Amadou Sidibé, Hannatou Souleymane Abarry, Sandrine Busière, Marine Vignon, Franck Lamontagne, Valery Ridde, Valériane Leroy, AIRE Research Study Group

## Abstract

**Background:** Despite the Integrated Management of Childhood Illness (IMCI) implemented at primary health centres (PHC) level, under-5 mortality remains high in sub-Saharan Africa. To improve the diagnosis and management of severe hypoxaemia, the AIRE project implemented the systematic use of pulse oximeters (PO) into IMCI consultations in PHCs in Burkina Faso, Guinea, Mali and Niger. We described the intervention sites, and measure their capacity to offer IMCI-care prior to project implementation.

**Methods:** A cross-sectional quantitative survey was conducted in all the PHCs and their district hospital (DH) of the AIRE project from March to July 2020.

**Results:** Overall, 215 PHCs and 8 DH were surveyed. Almost all the PHCs were public structures mainly managed by nurses. The IMCI strategy was in force in all PHCs with at least one IMCI-trained agent in more than 99% of the PHCs. At baseline, PO was available in only 2/215 (1%) PHCs and 4/8 (50%) DH. Overall, 35/215 (16%) PHCs have functional ambulance for managing referrals to DH, including two with mobile oxygen. IMCI consultations were free-of-fees in Burkina Faso and Niger, but charged for in Guinea and Mali (from US$0.5 to US$1). All the DH had capacities to provide specialised paediatric care for children under-5 years of age, although the use of PO was not systematic. Oxygen was available at all DH except one. Parents of children requiring hospitalisation had to pay out of pocket costs ranging from US$1.7 to US$8.4 per day.

**Conclusions:** This study revealed the absence of POs at PHC level and their low use at hospital level, as well as difficulties in managing referral to hospital of severe cases and access to mobile oxygen. It has guided the reasoned choice of the AIRE research sites, and the upgrading of PHCs including IMCI training before the project implementation.

**Study registration number:** PACTR202206525204526 registered on 06/15/2022.

**• What is already known on this topic:**

- Globally, the weakness of the healthcare system in West Africa had been demonstrated.
- In our knowledge, it is the first study in these countries that aimed to assess the capacity of health facilities to provide health care to ill children under-five. Few studies have provided some indicators, such as human resources or accessibility challenges, but not a real assessment.

**• What this study adds:** This study describes the weakness of the healthcare system in the four countries where the AIRE project has been implemented (Burkina Faso, Guinea, Mali, Niger), with:

- a shortage of skilled human resources in health, a lack of medical equipment, including Pulse Oximeters at primary healthcare centres, and their poor use at hospital level, and frequent shortages of essential medicines,
- financial problems in benefiting from health services, despite the total and partial exemption policies in force, which were not being properly applied,
- difficulties in organizing hospital transfers for severely ill children and the unavailability of oxygen during hospital transfers.

**• How this study might affect research, practice or policy:** The challenges identified through this study

- guided the upgrading of sites before the AIRE project implementation,
- raised awareness among health authorities of the many gaps in health systems that remain unresolved, especially the problem of hospital transfers and access to medical oxygen.

## INTRODUCTION

Children continue to face wide regional and income disparities in their chances of survival. Sub-Saharan Africa remains the region with the highest under-5 mortality rate in the world - 76 deaths per 1,000 live births in 2017 (1). In 2019, 1 in 13 children in sub-Saharan Africa died before reaching their fifth birthday - a risk 15 times higher than that of children born in high-income countries. Child mortality remains high despite progresses in child health management with the WHO Integrated Management of Childhood Illness (IMCI) guidelines at primary health centres (PHC) (2–4). Among the leading causes of death worldwide in 2017, pneumonia was estimated to account for 24% of deaths, followed by diarrhoea (15%) and malaria (9%) (1). In the West African region, neonatal conditions are more likely to be the leading cause of death (24%), followed by lower respiratory tract infections (16%), malaria (14.8%) and diarrhoea (13%) (5).

Hypoxemia is defined as a low haemoglobin oxygen saturation of less than 94% at sea level, as measured by Pulse Oximeter (PO) (peripheral oxygen saturation, SpO_2_) (6). The normal value of SpO_2_ at sea level ranges between 94% and 100%, needing adjustment at high altitude (> 2500 m) (7). Severe hypoxemia (SpO_2_<90%) is a well-known complication of pneumonia (8,9). It is a common sign of severity in children with acute respiratory illnesses, or with non-respiratory illnesses, significantly increasing their risk of death and requiring immediate oxygen, but remains under-diagnosed clinically (8,10). According to contexts, it is estimated that severe hypoxemia prevalence varies between 2% (9) and 80% (8,10,11) in children with severe illnesses.

The AIRE (Améliorer l’Identification des détresses Respiratoires chez l’Enfant) project aimed at improving the identification of respiratory distress in children under-5 years in West Africa. Funded by UNITAID, it was implemented since 2019 by a consortium of three NGOs (ALIMA, SOLTHIS, Terre des hommes (Tdh)) and the French Institute of Health and Medical Research (Inserm), with the support of PACCI program (Programme National de Lutte contre le Sida-French ANRS, and Coopération Nord-Sud, Côte d’Ivoire), and IRD (Institute of Research and Development) in four West African countries (Burkina Faso, Guinea, Mali, Niger). From 2020 to 2022, the research component was implemented in collaboration with the Ministries of Health (MoH), to assess the routine PO use integrated in the IMCI strategy for children under-5 in PHCs, in two health districts per country. It was expected that this intervention would improve the identification of severe hypoxemia in children under-5 at PHC for appropriate care management to improve their survival. Thus, a better knowledge of the baseline context at the country level was needed in terms of organisation and functioning of each national health care system and health policy, and the location and functioning of PHCs and their relative hospitals referral to guide and adapt the implementation of the AIRE project. This baseline assessment was specifically aimed to: i) describe the health context of the intervention health districts, ii) measure the capacity of health infrastructures to deliver IMCI to children under-5 (training, medical equipment, drugs and consumables) and to manage hospital referrals, iii) describe the information system of the PHCs and hospitals. iv) guide the selection of the AIRE research PHCs in each country to conduct the research component.

## METHODS

### Study design

A descriptive cross-sectional baseline survey was conducted in all the health structures (PHCs and their referral hospitals) covering the two health districts per country (Table 1) that were selected in the four countries to implement the AIRE project. We use the term district hospital (DH) for the referral hospitals in the district, even if they are not formally district hospitals.

**Table 1:** Distribution by country and district of the 215 Primary Health Centres (PHCs), and eight district hospitals (DH) included in the baseline site assessment of the AIRE project, March 2020-July 2020.

| Country | Period of survey | Health district | NGO implementer | Number of PHCs surveyed | Number of DH surveyed |
| --- | --- | --- | --- | --- | --- |
| Burkina Faso | March 12 to May 29, 2020 | Dédougou | Terre des hommes | 49 | 1 |
|  |  | Boromo | Terre des hommes | 38 | 1 |
| Guinea | March 16 to June 17, 2020 | Matoto | ALIMA | 16 | 1 |
|  |  | Télimélé | ALIMA | 14 | 1 |
| Mali | March 16 to March 26, 2020 | Dioila | ALIMA | 25 | 1 |
|  |  | Markala | Terre des hommes | 19 | 1 |
| Niger | March 16 to July 2020 | Niamey-IV | SOLTHIS | 12 | 1 |
|  |  | Dosso | SOLTHIS | 42 | 1 |
| Total |  |  |  | 215 | 8 |

### Study context

The four West African countries were chosen because of their high severe disease-related mortality, the weakness of their health care system, and the pre-existing involvement of the NGOs partners acting with the respective MoHs in supporting health care system activities and capacities. This choice increased the feasibility, and reduced the cost and risks (including the security perspective) of implementing the AIRE project. In each of the four countries, the project intervention took place in two health districts (Figure 1). In Burkina Faso, Tdh was leading the implementation activities, in Guinea, it was ALIMA, in Mali, ALIMA was in charge in the district of Dioila and Tdh in the district of Markala, and in Niger, it was Solthis.

**Figure 1:**
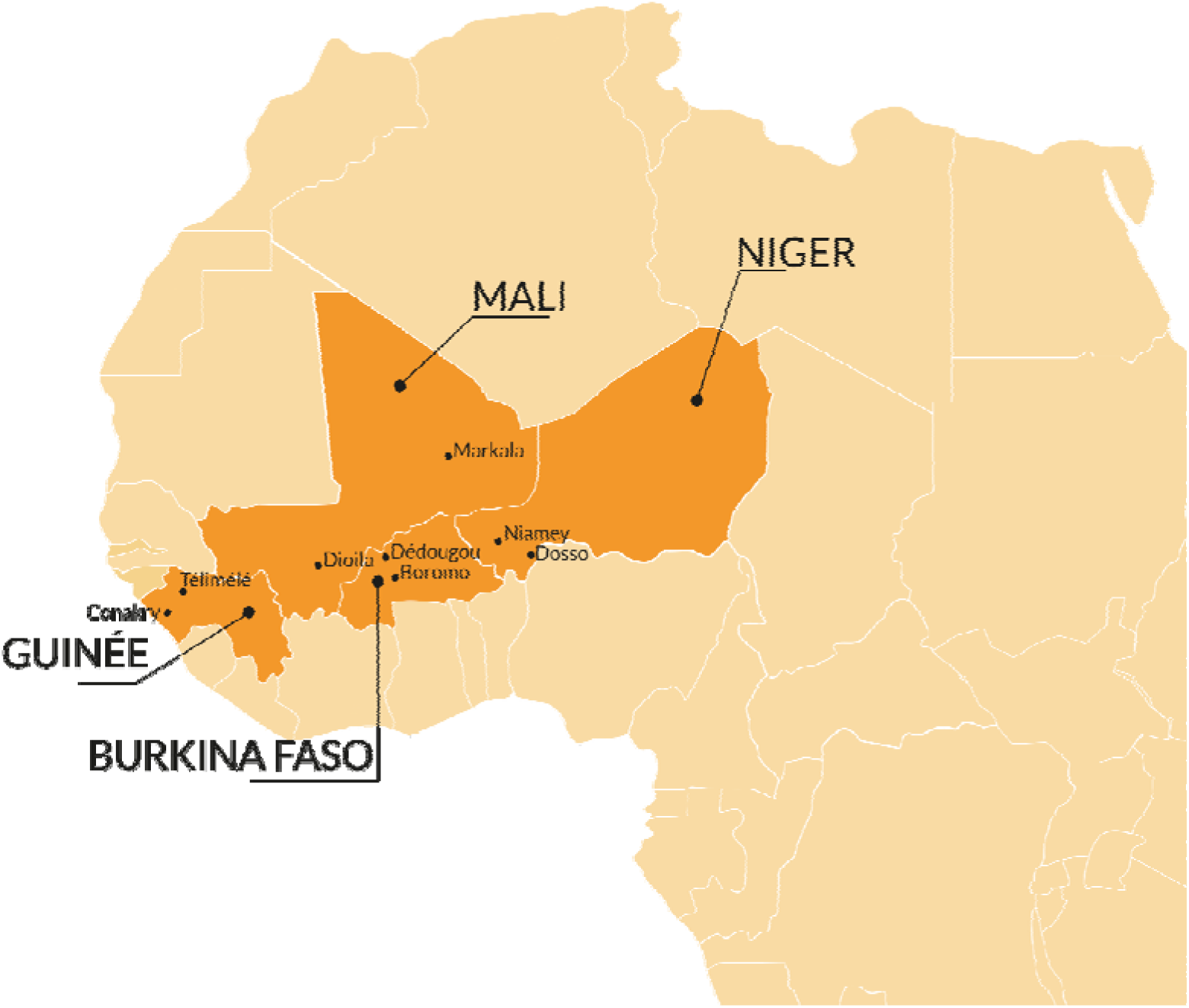
AIRE project implementation countries with area of intervention (2020–2022)

The four country’s health care systems are similarly organised in a pyramidal form at three levels: the health district at the bottom of the pyramid, the health region at the intermediate level and the central level at the top. The health facilities also integrate the same organization with at the first level, the peripheral health structures with health boxes or dispensaries followed by the health posts or PHC, and the DH at the health district level, then the regional hospitals at the intermediate level which are health reference structures for the district level and finally the national reference hospitals at the central level.

The IMCI strategy proposed by the WHO (2–4) for the care management of children under-5 years of age was in force In the four countries. This strategy is used at PHC level to classify children into three categories according to their illness severity (simple, moderate and severe), and to the age of the child in order to guide the appropriate treatment. It has been slightly adapted at each country level. In Burkina Faso and in the Markala health district in Mali, Tdh’s Integrated e-Diagnostic Approach (IeDA)(12) has digitized the IMCI guidelines and the health care workers (HCW) used it with a tablet. In the other health districts, HCW used paper-based IMCI.

Referral of seriously ill children identified at the PHC level is generally handled by the health system with PHC ambulance, or via a local organization, but can also be handled by regional or national ambulances.

Most West African countries have opted for total or partial free health care for children under-5. Care payment exemption policy was full in Burkina Faso and Niger for the care of all IMCI children (13,14). In Mali (15,16) and Guinea (17), the payment exemption policy was partial and limited to the treatment of four diseases: malaria, tuberculosis, HIV and malnutrition. In each country, these payment exemption policies apply at hospital level, but do not clearly apply to hospital referrals.

### Description of the AIRE health districts selected by country (Table 1)

In Burkina Faso, the two rural health districts selected were Boromo and Dédougou, among the six districts of the “Boucle du Mouhoun” region.

Boromo is equidistant (180 kms) between the capital Ouagadougou and the second city Bobo-Dioulasso. It has an area of 4,583 km^2^, covering 13% of the Boucle du Mouhoun region. The updated data from the general census of the human population in 2019, gave a population of 305,193 inhabitants and an estimated density of 67 inhabitants per km². The district accounted 38 PHCs, one medical centre and one medical centre with a surgical unit as the DH. In this district, 68% of the population lived within 5 kms from a health facility while 12% lived beyond 10 kms. At the DH, a gynaecologist was the single specialist, with general practitioners and specialised nurses. Natural barriers make accessibility to care difficult during the rainy season occurring yearly from May to October.
Dédougou health district covers an area of 1,354 km^2^. The population of the district was estimated at 424,240 in 2019. The health district has 49 public PHCs, 7 private PHCs and 1 DH. The proportion of those living in rural areas is 88%; 59% of the population lives within 5 kms from a health facility; and 12% live beyond more than 10 kms. The DH of Dédougou is located in the capital of the “Boucle du Mouhoun” region covers the entire region. It is the second level of reference for the health district of Dédougou. The medical staff is mainly composed of general practitioners, and few specialists such as obstetricians, paediatricians and specialized nurses.

In Guinea, the two health districts selected were Matoto and Télimélé.

Matoto is one of the five cities that constitute the city of Conakry, the capital of Guinea, which is a peninsula 34 kms long and 1 to 6 kms wide. Matoto health district has a total of 16 PHC (7 public and 9 private). It is important to note the existence of small private practices that are not included in the local health system. The total population of the district in 2019 was 778,655. The Matoto facilities refer either to the Matam hospital (DH) or directly to the INSE (Institut de nutrition et de santé de l’enfant), Donka or the Ignace Deen hospitals, as appropriate. The INSE and Donka hospitals are in the same location. INSE receives paediatric malnutrition cases and has a specific neonatology unit. The paediatric ward in Donka treats other paediatric cases.
The health district of Télimélé is a rural and mountainous district. It has a total of 14 health areas for the 14 communes (one urban and 13 rural). The population was 178,998 in 2019 (18) with a density of 36 inhabitants per km^2^. Thirty percent of the population of Telimélé lived within less than 5 kms from the health facility and the rest of the population beyond 10 kms. Distances between the PHCs and the DH are significant and the roads difficult to use due to the lack of adequate means of transport and population poverty, which complicates hospital referral.

In Mali, the project was implemented in the health districts of Dioila and Markala.

The cercle of Dioila became a health region in 2018, located in the centre of southern Mali. The health district covers 24 health areas for 355 villages, with a population of 687,160 inhabitants in 2018. Forty percent of the population is located within a 5kms distance from a PHC and 86% within 15kms. Distances between some PHC and the DH are considerable with impracticable roads that are sometimes even cut off during the rainy season, which makes referral hospital extremely difficult.
Markala has a total of 19 health zones for 10 rural municipalities. The population is 342,398. Sixty-five percent of the population lives within 5 kms of health structure and 35% beyond 15 kms. Distances between the PHC and the DH are important and roads difficult to use due to the absence of adequate means of transport and the poverty of the populations, which makes also referrals difficult.

In Niger, the two health districts selected were Dosso and Niamey-IV.

The Regional Hospital Centre of Niamey (CHRN) is the referral hospital of the Niamey-IV’s PHCs based in an urban area. Its paediatric service accounted five hospitalization wards with a total of 82 beds. This hospital is within the closest distance compared to other AIRE DHs. However, it should be noted that during peak period, each bed can accommodate three children.
In Dosso, a rural area, the Mother and Child Health Centre (CSME) of the Dosso Health District received referrals from children and mothers. There were two paediatric wards; with 42 beds and 105 beds.

### Data collection

From March 2020 to July 2020, two standardised questionnaires were specifically conceived, one for the PHCs and another one for the DH, based on the WHO standards for improving quality of maternal, newborn, children and young adolescents care in health facilities (19,20). Each questionnaire was completed based on the healthcare facility managers’ declarations, and the health care activities registers and reports. The data collected by the research study team were mainly: geographical location, accessibility to PHCs for the population and research teams (distance, road practicability, isolation during the rainy season, and insecurity), human resources, infrastructures, attendance of IMCI consultations for children under-5, available medical and technical equipment and health care services, availability of PO and basic medicines including antibiotics and oxygen (oxygen concentrators and/or bottled oxygen), cost of services, health data management tools, organisation of referrals, etc. Data were collected using an electronic questionnaire installed on tablets using Kobocollect software. This study was conducted with the approval from the four MoHs in 2020.

### PHCs selection as research sites

This baseline survey has guided the selection of the four PHCs per country (two per health district) used as the AIRE research sites and dedicated to the further individual data collection planned in the research protocol (21). This selection was based on the following main criteria: the PHC had to i) have adopted the IMCI approach for the care of children under-5, ii) not previously introduce PO for the care of children, iii) be accessible to the members of the research team, iv) have available internet access, v) have a minimal organisation system for hospital transfer of children, vi) not have been selected to carry out the specific Covid-19 interventions. Among the remaining eligible PHCs, based on 2019 data from each health facility, we selected at least one large and one medium/small PHC per health district on the basis of yearly IMCI attendance (small PHCs: ≤1000, medium PHCs: [1000-3000], large PHCs >3000 IMCI consultations/year) to be representative of PHC attendance when studying the implementation process of AIRE intervention. Finally, the preselected PHCs shortlisted were presented to the MoH’ authorities for approval before obtaining any ethical clearance for the AIRE research project.

### Data analysis

A narrative description of the characteristics of the 215 PHCs and eight DH was completed, then for the 16 PHCs selected for the research, with quantitative data analysed using means and standard deviations or median and interquartile ranges. Proportions were used to describe categorical data. The data analysis was carried out with R software version 3.6.2.

### Patient and public involvement

This study was conducted using programmatic data collected routinely with each MOH authorization. No patient was involved in the study. Patients did not contribute to the writing or editing of this manuscript.

## RESULTS

### Characteristics of the 215 Primary Health Centres of the AIRE project (Table 2)

Overall, 93% of the PHCs were public sites. They are all public in Burkina Faso, Télimélé (Guinea), Markala (Mali) and Dosso (Niger). However, 63% of the PHCs in Matoto (Guinea), 33% in Niamey-IV (Niger) and 4% in Dioila (Mali) were private or confessional. Overall, 83% were rural PHCs; the urban PHCs were Matoto and Niamey-IV health district. The accessibility to the PHCs for study teams in terms of geographical barriers (impassable roads, hill or mountains, waterways…) or insecurity, varied from 14% in Télimélé to 69% in Matoto. More than 95% of the PHCs were covered by a telephone network and more than 80% of PHCs in each health districts had internet access with the exception of the Boromo district (61% of PHCs) in Burkina Faso. Overall, 76% of the AIRE PHCs were small to medium sized centers in terms of IMCI attendance while high attendance was observed in the Niamey district with 8/12 (67%) large PHCs.

**Table 2:**
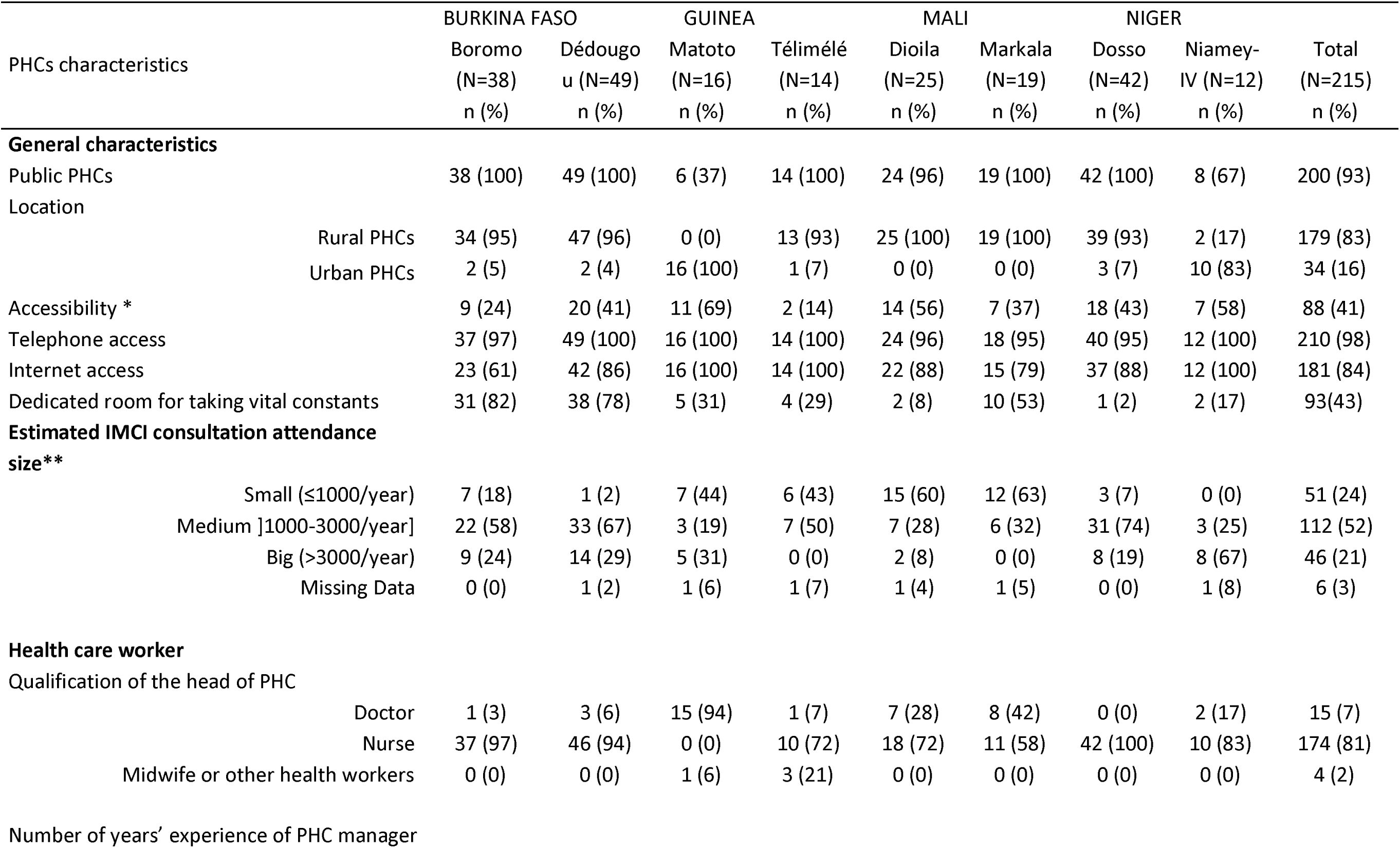
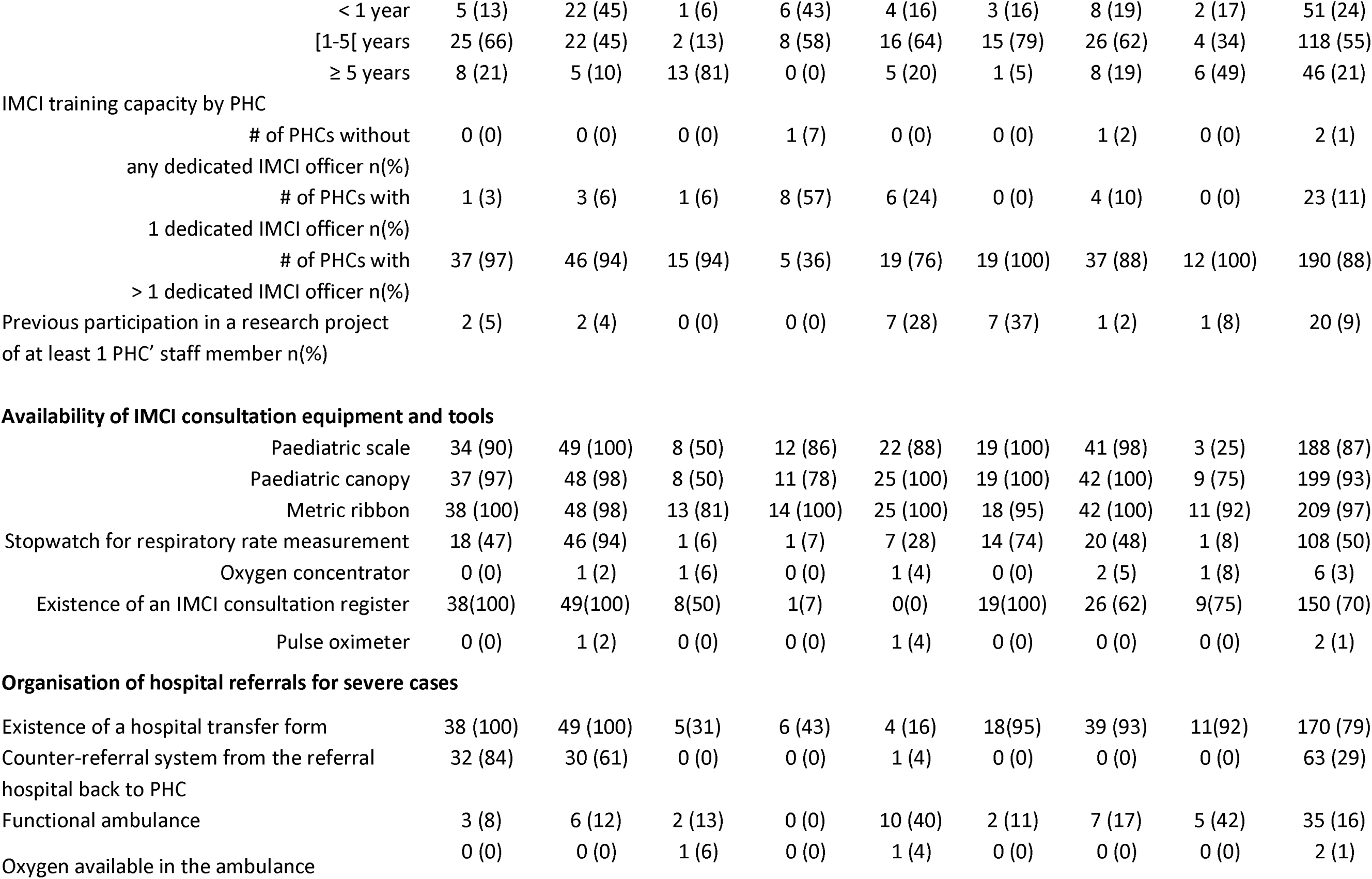
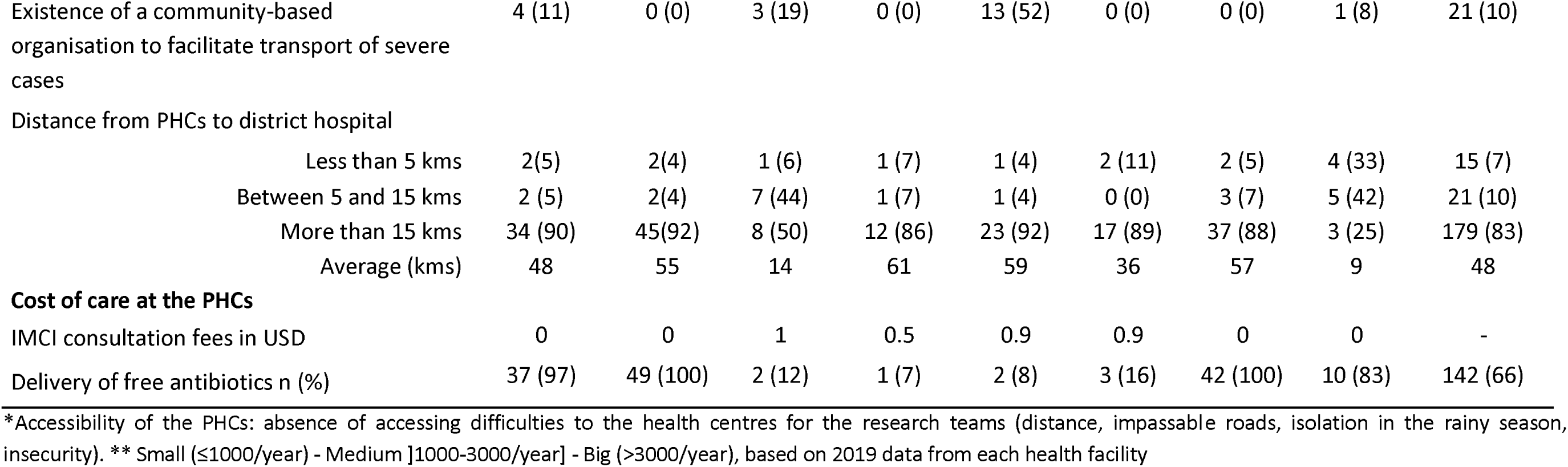
Characteristics of the 215 PHCs involved in the AIRE project by health district and country, March-July 2020.

|  |  | BURKINA FASO |  | GUINEA |  | MALI |  | NIGER |  |  |
| --- | --- | --- | --- | --- | --- | --- | --- | --- | --- | --- |
| PHCs characteristics |  | Boromo<br>(N=38)<br>n (%) | Dédougou<br>(N=49)<br>n (%) | Matoto<br>(N=16)<br>n (%) | Télimélé<br>(N=14)<br>n (%) | Dioila<br>(N=25)<br>n (%) | Markala<br>(N=19)<br>n (%) | Dosso<br>(N=42)<br>n (%) | Niamey-IV (N=12)<br>n (%) | Total<br>(N=215)<br>n (%) |
| **General characteristics** |  |  |  |  |  |  |  |  |  |  |
| Public PHCs |  | 38 (100) | 49 (100) | 6 (37) | 14 (100) | 24 (96) | 19 (100) | 42 (100) | 8 (67) | 200 (93) |
| Location |  |  |  |  |  |  |  |  |  |  |
|  | Rural PHCs | 34 (95) | 47 (96) | 0 (0) | 13 (93) | 25 (100) | 19 (100) | 39 (93) | 2 (17) | 179 (83) |
|  | Urban PHCs | 2 (5) | 2 (4) | 16 (100) | 1 (7) | 0 (0) | 0 (0) | 3 (7) | 10 (83) | 34 (16) |
| Accessibility * |  | 9 (24) | 20 (41) | 11 (69) | 2 (14) | 14 (56) | 7 (37) | 18 (43) | 7 (58) | 88 (41) |
| Telephone access |  | 37 (97) | 49 (100) | 16 (100) | 14 (100) | 24 (96) | 18 (95) | 40 (95) | 12 (100) | 210 (98) |
| Internet access |  | 23 (61) | 42 (86) | 16 (100) | 14 (100) | 22 (88) | 15 (79) | 37 (88) | 12 (100) | 181 (84) |
| Dedicated room for taking vital constants |  | 31 (82) | 38 (78) | 5 (31) | 4 (29) | 2 (8) | 10 (53) | 1 (2) | 2 (17) | 93(43) |
| **Estimated IMCI consultation attendance size\*\*** |  |  |  |  |  |  |  |  |  |  |
| | Small ($\leq 1000$/year) | 7 (18) | 1 (2) | 7 (44) | 6 (43) | 15 (60) | 12 (63) | 3 (7) | 0 (0) | 51 (24) |
|  | Medium ]1000-3000/year] | 22 (58) | 33 (67) | 3 (19) | 7 (50) | 7 (28) | 6 (32) | 31 (74) | 3 (25) | 112 (52) |
|  | Big (>3000/year) | 9 (24) | 14 (29) | 5 (31) | 0 (0) | 2 (8) | 0 (0) | 8 (19) | 8 (67) | 46 (21) |
|  | Missing Data | 0 (0) | 1 (2) | 1 (6) | 1 (7) | 1 (4) | 1 (5) | 0 (0) | 1 (8) | 6 (3) |
| **Health care worker** |  |  |  |  |  |  |  |  |  |  |
| Qualification of the head of PHC |  |  |  |  |  |  |  |  |  |  |
|  | Doctor | 1 (3) | 3 (6) | 15 (94) | 1 (7) | 7 (28) | 8 (42) | 0 (0) | 2 (17) | 15 (7) |
|  | Nurse | 37 (97) | 46 (94) | 0 (0) | 10 (72) | 18 (72) | 11 (58) | 42 (100) | 10 (83) | 174 (81) |
|  | Midwife or other health workers | 0 (0) | 0 (0) | 1 (6) | 3 (21) | 0 (0) | 0 (0) | 0 (0) | 0 (0) | 4 (2) |
| Number of years' experience of PHC manager |  |  |  |  |  |  |  |  |  |  |

|  |  |  |  |  |  |  |  |  |  |
| --- | --- | --- | --- | --- | --- | --- | --- | --- | --- |
| < 1 year | 5 (13) | 22 (45) | 1 (6) | 6 (43) | 4 (16) | 3 (16) | 8 (19) | 2 (17) | 51 (24) |
| [1-5[ years | 25 (66) | 22 (45) | 2 (13) | 8 (58) | 16 (64) | 15 (79) | 26 (62) | 4 (34) | 118 (55) |
| ≥ 5 years | 8 (21) | 5 (10) | 13 (81) | 0 (0) | 5 (20) | 1 (5) | 8 (19) | 6 (49) | 46 (21) |
| **IMCI training capacity by PHC** |  |  |  |  |  |  |  |  |  |
| # of PHCs without any dedicated IMCI officer n(%) | 0 (0) | 0 (0) | 0 (0) | 1 (7) | 0 (0) | 0 (0) | 1 (2) | 0 (0) | 2 (1) |
| # of PHCs with 1 dedicated IMCI officer n(%) | 1 (3) | 3 (6) | 1 (6) | 8 (57) | 6 (24) | 0 (0) | 4 (10) | 0 (0) | 23 (11) |
| # of PHCs with > 1 dedicated IMCI officer n(%) | 37 (97) | 46 (94) | 15 (94) | 5 (36) | 19 (76) | 19 (100) | 37 (88) | 12 (100) | 190 (88) |
| Previous participation in a research project of at least 1 PHC' staff member n(%) | 2 (5) | 2 (4) | 0 (0) | 0 (0) | 7 (28) | 7 (37) | 1 (2) | 1 (8) | 20 (9) |
| **Availability of IMCI consultation equipment and tools** |  |  |  |  |  |  |  |  |  |
| Paediatric scale | 34 (90) | 49 (100) | 8 (50) | 12 (86) | 22 (88) | 19 (100) | 41 (98) | 3 (25) | 188 (87) |
| Paediatric canopy | 37 (97) | 48 (98) | 8 (50) | 11 (78) | 25 (100) | 19 (100) | 42 (100) | 9 (75) | 199 (93) |
| Metric ribbon | 38 (100) | 48 (98) | 13 (81) | 14 (100) | 25 (100) | 18 (95) | 42 (100) | 11 (92) | 209 (97) |
| Stopwatch for respiratory rate measurement | 18 (47) | 46 (94) | 1 (6) | 1 (7) | 7 (28) | 14 (74) | 20 (48) | 1 (8) | 108 (50) |
| Oxygen concentrator | 0 (0) | 1 (2) | 1 (6) | 0 (0) | 1 (4) | 0 (0) | 2 (5) | 1 (8) | 6 (3) |
| Existence of an IMCI consultation register | 38(100) | 49(100) | 8(50) | 1(7) | 0(0) | 19(100) | 26 (62) | 9(75) | 150 (70) |
| Pulse oximeter | 0 (0) | 1 (2) | 0 (0) | 0 (0) | 1 (4) | 0 (0) | 0 (0) | 0 (0) | 2 (1) |
| **Organisation of hospital referrals for severe cases** |  |  |  |  |  |  |  |  |  |
| Existence of a hospital transfer form | 38 (100) | 49 (100) | 5(31) | 6 (43) | 4 (16) | 18(95) | 39 (93) | 11(92) | 170 (79) |
| Counter-referral system from the referral hospital back to PHC | 32 (84) | 30 (61) | 0 (0) | 0 (0) | 1 (4) | 0 (0) | 0 (0) | 0 (0) | 63 (29) |
| Functional ambulance | 3 (8) | 6 (12) | 2 (13) | 0 (0) | 10 (40) | 2 (11) | 7 (17) | 5 (42) | 35 (16) |
| Oxygen available in the ambulance | 0 (0) | 0 (0) | 1 (6) | 0 (0) | 1 (4) | 0 (0) | 0 (0) | 0 (0) | 2 (1) |
| Existence of a community-based organisation to facilitate transport of severe cases | 4 (11) | 0 (0) | 3 (19) | 0 (0) | 13 (52) | 0 (0) | 0 (0) | 1 (8) | 21 (10) |
| Distance from PHCs to district hospital |  |  |  |  |  |  |  |  |  |
| Less than 5 kms | 2(5) | 2(4) | 1 (6) | 1 (7) | 1 (4) | 2 (11) | 2 (5) | 4 (33) | 15 (7) |
| Between 5 and 15 kms | 2 (5) | 2(4) | 7 (44) | 1 (7) | 1 (4) | 0 (0) | 3 (7) | 5 (42) | 21 (10) |
| More than 15 kms | 34 (90) | 45(92) | 8 (50) | 12 (86) | 23 (92) | 17 (89) | 37 (88) | 3 (25) | 179 (83) |
| Average (kms) | 48 | 55 | 14 | 61 | 59 | 36 | 57 | 9 | 48 |
| **Cost of care at the PHCs** |  |  |  |  |  |  |  |  |  |
| IMCI consultation fees in USD | 0 | 0 | 1 | 0.5 | 0.9 | 0.9 | 0 | 0 | - |
| Delivery of free antibiotics n (%) | 37 (97) | 49 (100) | 2 (12) | 1 (7) | 2 (8) | 3 (16) | 42 (100) | 10 (83) | 142 (66) |

\*Accessibility of the PHCs: absence of accessing difficulties to the health centres for the research teams (distance, impassable roads, isolation in the rainy season, insecurity). \*\* Small ( $\leq 1000$ /year) - Medium ]1000-3000/year] - Big ( $> 3000$ /year), based on 2019 data from each health facility

### PHCs health staff: qualification and seniority of the PHCs manager, coverage of IMCI trained HCW

Most of the PHCs (81%), varying from 58% to 100% according to health districts) were managed by nurses, except in the Matoto district where management was entrusted to the doctor. The seniority of these PHC managers varied from 0 to 5 years, except in Matoto where more than 80% had been in charge for more than 5 years. All the PHCs have at least one HCW trained to IMCI guidelines apart from one PHC in the health districts of Dosso and Télimélé.

### Medical equipment including PO and IMCI consultation registers

The medical equipment and registers needed to conduct an IMCI consultation were generally absent in the health facilities. One out of four (25%) of the PHCs in Niamey-IV had a paediatric scale, in Matoto, 1 out of 2 PHCs had a paediatric scale and only one out of 16 had a stopwatch. Among the 215 PHCs, only 2 PHCs (1 in Dédougou and 1 in Dioïla) had a PO. Oxygen concentrator was scarce, 6/215 (3%) PHCs with only one in Burkina Faso (Dédougou), Guinea (Matoto), Mali (Dioïla) and three in Niger (one in Niamey-IV and two in Dosso). Use of IMCI consultation registers were almost inexisting in Télimélé (Guinea) and Dioïla (Mali) but generally available in the other AIRE DH. In the 137 PHCs (64%) where the IMCI consultation register was available, it was not completed correctly. Of note, the main difference in IMCI classification reference between countries was the chest indrawing, considered as a danger sign only in Guinea.

### Management of severe IMCI cases: hospital referral

To refer children with serious illness, hospital referral forms were available in all PHCs, except in Matoto (31% of PHCs), Dioila (43% of PHCs), and Telimele (16% of PHCs). Overall, no counter-referral system to inform the original PHC of the outcome of referred patients was in place, except in Burkina Faso. Functional ambulances were available in only 35 of the 215 PHCs (16%) covering all the heath districts, but none in the Télimélé district. Only two (1%) PHCs (one in Dioïla and one in Matoto) could deliver oxygen during the transfer to hospital. Depending on the health district, 9% (21/215) of the PHCs have established community-based initiatives to organise referral of severe cases to the DH but it was distributed only among the districts of Boromo, Matoto, Dioila and Niamey-IV. Finally, in terms of distances between PHCs and DH, only half of the PHCs in Matoto and 85% in Niamey-IV were less than 15 kms from the DH. In all the other districts, more than 86% of the PHCs are located at more than 15 kms from the DH, ranging from 36 kms in Markala (Mali) to 61 kms in Télimélé. The average distance between PHC and their DH varied widely among health district from 4.5 kms (Niamey-IV) to 96.5 kms (Dosso).

### Cost of care

At the time of launching the AIRE project, IMCI consultations at the PHCs were free of charge in Burkina Faso and Niger, but charged in Guinea and Mali with a declared prices varying from US$0.5 to US$1, respectively. The provision of free antibiotics was possible in the PHCs of Burkina Faso and Niger, but not in the other countries.

### Characteristics of the16 research sites selected for individual data collection (Table 3)

The 16 research sites selected with the MoH approval were public sites. Most of them were rural sites, with an average of three HCW trained to perform IMCI consultations (range:1-6). All have at least one consultation room where IMCI consultations can take place, and have access to Internet. None of them has yet introduced the PO in practice. The equipment needed for IMCI consultations was available in most of the PHCs except in two sites in Guinea (PHC Télimélé and PHC Santou) and one site in Niger (PHC Aéroport2) where paediatric scales, height measuring towers, metric tape and stopwatch were generally absent; these sites had been upgraded before the AIRE research started. In terms of capacity to manage the referral of severe cases identified at these research sites, 6 of the 16 PHCs (37.5%) have an ambulance: none in Guinea, one in Mali, two in Burkina Faso and three in Niger. But none of them was equipped to provide mobile oxygen for referral to the DH, which is 33.3 kms away from the research PHCs on average (min=5 kms; max=75 kms). Télimélé’ PHC were the exception because they are close to the DH. The research sites were generally geographically accessible and safe for the population, except for the Sibila PHC (Mali) and the Santou and Sarékaly sites (Guinea) due to impassable roads, especially in the rainy season, and the presence of rivers, hills and mountains. Otherwise, the research sites remain easily accessible for the project teams.

**Table 3:**
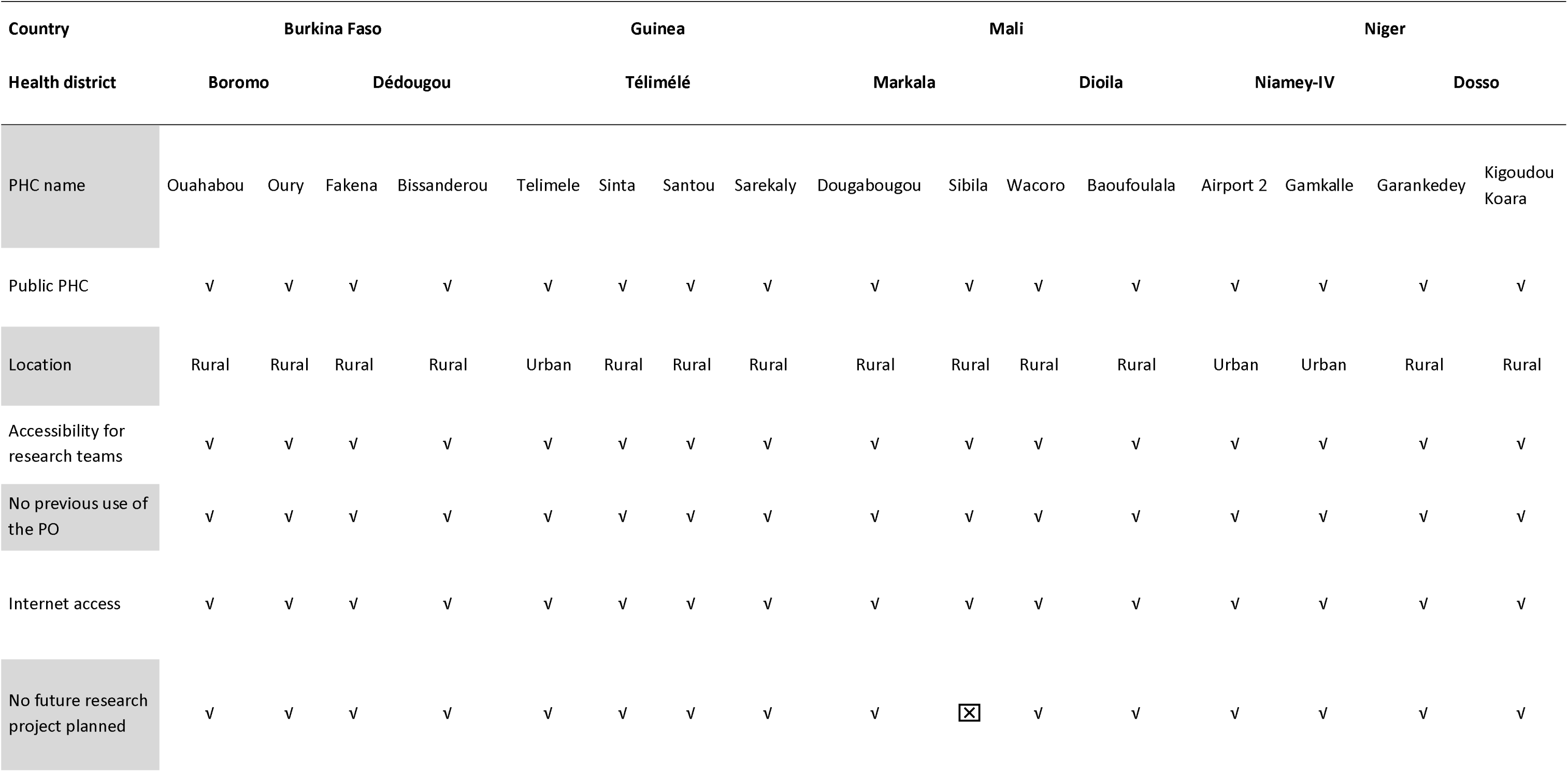
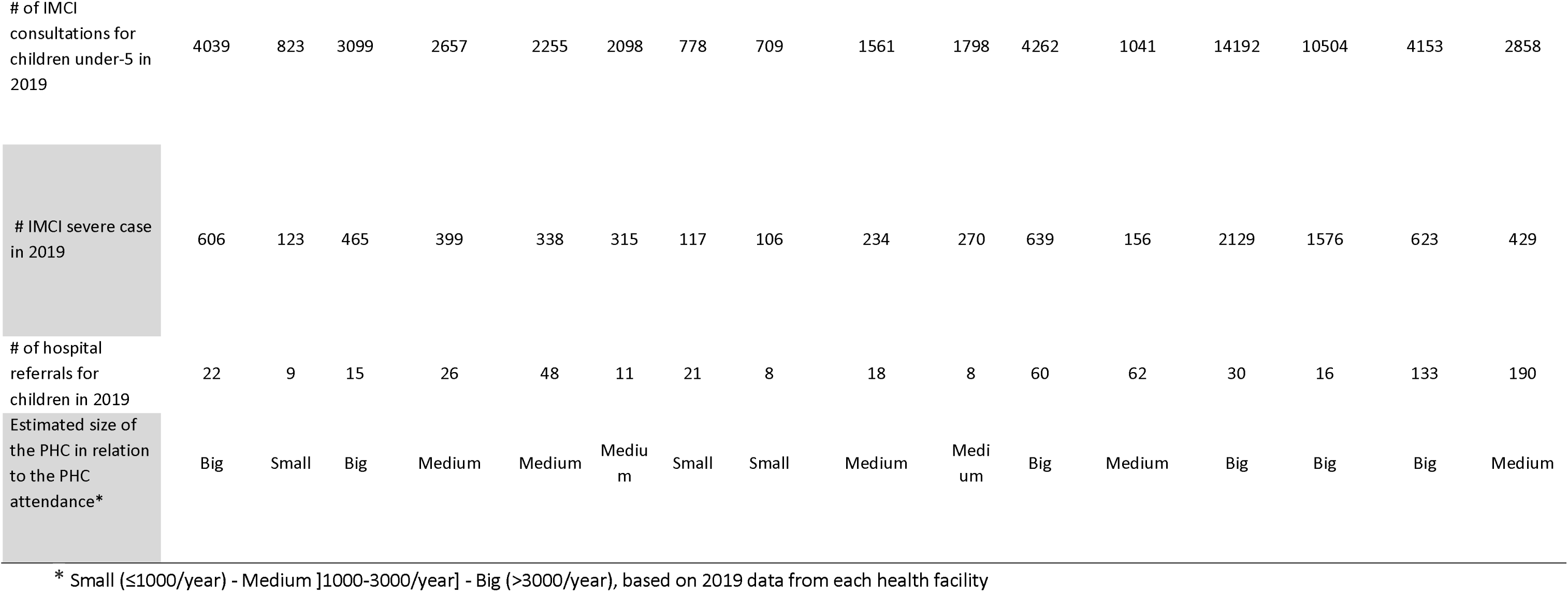
Description of the 16 AIRE research PHCs selected by district and country.

### Characteristics of DH in AIRE project countries (Table 4)

#### Geographical distribution, infrastructure in referral hospitals

Six of the eight hospitals are located in urban areas, except in Mali. All were able to deliver paediatric care to sick children. They have a general consultation and hospitalisation services, a nutritional recovery unit and a paediatric ward. However, paediatric emergencies did not exist in Télimélé (Guinea) and in Boromo (Burkina Faso); neonatology service did not exist in Guinea, nor at Markala and Boromo districts, respectively in Mali and Burkina Faso. All the AIRE DH are supplied with electricity, either from the national grid for all formal hospitals, or with alternative solutions such as generators or solar panels.

**Table 4:** Characteristics of the eight district hospitals involved in the AIRE project by health district and country, March-July 2020. (√ means Yes, ⌧ means absence)

| Characteristics of district hospitals | Burkina Faso |  | Guinea |  | Mali |  | Niger |  |
| --- | --- | --- | --- | --- | --- | --- | --- | --- |
|  | CMA of Boromo | CHR Dédougou | Ignace Deen Hospital | Télimélé Prefectural Hospital | Dioila Hospital | Markala Hospital | CSME of Dosso | CHRN of Niamey-IV |
| **Characteristics** |  |  |  |  |  |  |  |  |
| Urban area | ✓ | ✓ | ✓ | ✓ | ☒ | ☒ | ✓ | ✓ |
| **Care services offer** |  |  |  |  |  |  |  |  |
| Department of Paediatrics | ✓ | ✓ | ✓ | ✓ | ✓ | ✓ | ✓ | ✓ |
| Department of Neonatology | ☒ | ✓ | ☒ | ☒ | ✓ | ☒ | ✓ | ✓ |
| Paediatric emergencies | ☒ | ✓ | ✓ | ☒ | ✓ | ✓ | ✓ | ✓ |
| General consultations | ✓ | ✓ | ✓ | ✓ | ✓ | ✓ | ✓ | ✓ |
| Hospitalisation | ✓ | ✓ | ✓ | ✓ | ✓ | ✓ | ✓ | ✓ |
| Nutritional Recovery and Education | ✓ | ✓ | ✓ | ✓ | ✓ | ✓ | ✓ | ✓ |
| **Source of electrical energy supply** |  |  |  |  |  |  |  |  |
| National electricity grids | ✓ | ✓ | ✓ | ✓ | ✓ | ✓ | ✓ | ✓ |
| Generator set | ✓ | ☒ | ✓ | ☒ | ✓ | ✓ | ✓ | ✓ |
| Solar panels | ☒ | ☒ | ✓ | ☒ | ☒ | ☒ | ☒ | ☒ |
| **Characteristics of health workers** |  |  |  |  |  |  |  |  |
| Paediatricians (#/DH) | 0 | 4 | 5 | 0 | 0 | 0 | 4 | 5 |
| General practitioners (#/DH) | 1 | 4 | 5 | 2 | 5 | 3 | 0 | 1 |
| Nurses (#/DH) | 3 | 34 | 11 | 5 | 8 | 3 | 12 | 10 |
| Midwives (#/DH) | 0 | 9 | 0 | 7 | 3 | 7 | 1 | 5 |
| Biomedical technician | √ | √ | √ | √ | √ | ☒ | √ | √ |
| Staff training on oxygen therapy | ☒ | ☒ | √ | ☒ | √ | ☒ | √ | ☒ |
| **Availability and use of consultation facilities** |  |  |  |  |  |  |  |  |
| Functional paediatric scale | ☒ | √ | ☒ | √ | √ | √ | √ | √ |
| Availability of a measuring rod | √ | √ | ☒ | √ | √ | √ | √ | √ |
| Use of PO in the paediatric ward | √ | √ | ☒ | ☒ | √ | √ | ☒ | ☒ |
| Knowledge of the SpO<sub>2</sub> threshold used for oxygenation of a child under-5 years of age (<90%) | ☒ | ☒ | ☒ | ☒ | √ | ☒ | ☒ | ☒ |
| **Oxygen supply and delivery** |  |  |  |  |  |  |  |  |
| Oxygen availability | √ | √ | √ | ☒ | √ | √ | √ | √ |
| Means of oxygen distribution |  |  |  |  |  |  |  |  |
| Oxygen in cylinders | √ | √ | ☒ | ☒ | ☒ | √ | √ | √ |
| Wall circuit for the operating theatre | ☒ | ☒ | ☒ | ☒ | √ | ☒ | ☒ | ☒ |
| # functional oxygen concentrators (n) | 1 | 4 | 1 | 0 | 2 | 0 | 5 | 7 |
| Frequent breaks in oxygen | ☒ | √ | √ | √ | ☒ | √ | ☒ | ☒ |
| **Cost of care** |  |  |  |  |  |  |  |  |
| Consultation fees for children under-5 in US \$ | 0 | 0 | 0.5 | 1.1 | 1.7 | 0.9 | 0 | 0 |
| Daily hospitalisation fees US \$ | 1.7 | 0 | 6.3 | 8.4 | 2.6 | 0 | 0 | 0 |

#### Human resources at DH level

During the survey, the presence of several nurses (min=3; max=34 per DH) and at least one doctor (min=1; max=10 per DH) was noted. However, there were no paediatrician in the DH of Boromo (Burkina Faso), Télimélé (Guinea), Dioila and Markala (Mali). With regard to biomedical maintenance, all (7/8: 88%) but the Markala DH had a biomedical technician in place.

#### Availability of medical equipment for consultation, and use of the paediatric PO at DH level

The paediatric height meter (88%) and scale (75%) were tools generally available in the AIRE DH. No PO was used in the paediatric wards in Guinea and in Niger but it was available in 100% of paediatric wards in Burkina Faso and Mali. The SpO_2_ threshold indicating oxygen therapy for severely sick children (<90%) (6) was unknown to the clinicians for 87.5% of DH. Only the HCW from the Dioila referral hospital in Mali knew the correct threshold.

#### Availability and delivery of oxygen and antibiotic prescription and cost of care at DH level

Regarding the availability of oxygen, the prefectural hospital of Télimélé in Guinea had neither oxygen nor a distribution system in place at the time of the survey. In the other seven DHs, several means of oxygen distribution were available and balanced each other out. In all AIRE DH, the most prescribed antibiotics were Ceftriaxone for 63% of DH followed by Ampicillin for 25% of DH and Gentamicin for 12% of DH. The consultation for children under-5 years old was declared free of charge in the reference hospitals in Burkina Faso and Niger. In the other two countries, the declared prices of the consultation varied between US$0.5 and US$1.7. The daily prices of hospitalisation were US$0 in Niger, Markala in Mali and Dédougou in Burkina Faso, and varying from US$1.7 to US$8.4 in the other DHs.

## DISCUSSION

This assessment of the AIRE health facilities was conducted in 2020 at the beginning of the Covid-19 pandemic, before the start of the AIRE intervention and provide a picture of the context in which the project was implemented and will help in a better understanding of the future results.

Firstly, although poorly applied, most PHCs were actually able to provide IMCI consultations with at least one HCW trained per site. The PO was not introduced at the PHC level in the four AIRE countries, including in health districts where electronic IeDA-based IMCI support (12,22): it was only present in 1% of the PHCs and available in half of the DH, in paediatric wards. Other medical equipment often used for IMCI consultations of children under-5 years of age were not also always available at all the PHC and DH. Similar findings have been found reported in PHCs in Uganda and Malawi respectively (23,24). In Nigeria, Graham’s assessment revealed in 2021 that none of the 28 PHCs surveyed had a PO and all 3 hospitals had them but in only one (33%), it was functional (23). Despite the WHO indications in the 2014 IMCI guidelines which recommend the use of PO (if available) (3,4,25) for all the children under-5 with respiratory signs at the PHC level, it is noted that the countries of implementation have not actually implemented the recommendation to make PO available at this level of the health pyramid. This situation is widespread in West Africa but also in East Africa and Asia (26) and raised other problems such as the adequate supply of medical equipment including the PO to conduct IMCI consultations, but also the question of the quality or level of IMCI training of the HCW.

Secondly, our findings shown the weakness of the AIRE countries’ health system with insufficient health care resources. The lack of qualified HCW partially explain low access to health services, that was already described elsewhere (27–30). Our baseline survey specifically demonstrated the challenges in referring severely ill children to the hospital and for those hypoxemic, the inaccessibility to mobile oxygen during transfer to the district hospital. As a matter of fact, based on the 2019 referral estimation from the eight health district, only 7.9% (n=677) of IMCI severely ill children identified at the PHC level were referred to the hospital, a significantly lower proportion than the expected 15% estimated by Floyd(8). Even though oxygen was available in almost all AIRE DH, it was noted a lack of an oxygen delivery system at the PHC level (only 3%) and lack of ambulance (only 1%) able to administer oxygen therapy. In an assessment in Nigeria, H. Graham showed that 43% of PHCs (12/28) had an oxygen source (23). At the tertiary hospital level, the situation was improved with 100% (3/3) having access to oxygen. Oxygen distribution circuits are not available at the PHC level but rather at the hospital level. In reality, health policies in West Africa are not oriented towards oxygen administration at the PHC level where hypoxemia is under-diagnosed in the absence of PO use(31–34). But it could make sense to invest in oxygen delivery to PHCs if hypoxemic children are better diagnosed using PO. However, several parameters will need to be considered, including the quality of training for HCW, and the need for scientific evidence and assurance that it will be cost-effective. Another option would be to define a more sensitive threshold of SpO_2_ than the 90%, for indicating hospital referral to take into account the delay between the identification and the oxygen therapy corresponding to the hospital referral and care management with adequate referral systems to enable cases of hypoxaemia to arrive at the hospital in time to benefit from oxygen therapy. Indeed, our survey revealed the absence of an organised referral system in most of the PHCs, with no functional ambulance available and none with mobile oxygen on board. In addition, very few PHCs (10%) had a community structure involved in organizing the referral of severe cases. We noted that despite several initiatives such as the implementation of the additional cent in Niger (35) or community-based organisations in Mali or Burkina Faso(36), the field reality is different; hospital transfers are unaffordable for most of the families, and not effectively functional. This is a weak link in the chain of care for hypoxemic children in the four countries. In brief, the question of hospital transfers of children with serious illnesses remains unresolved with the scarcity of functional ambulances and community initiatives.

Finally, concerning the costs, in Niger and Burkina Faso, the baseline assessment found that parents should not have to pay for consultations at the PHC or at the DH in general (except in Boromo DH). There were no hospitalization costs declared in DH either, but in Mali and Guinea, families were expected to pay fees ranging from US$0.5 to US$1.0 at the PHCs level to US$1.7 at the DH for consultation and daily hospital charges ranging from US$2.6 to US$8.4. This is a picture of the policies of free healthcare for children under-5 implemented by governments in each of the four project countries. It should be noted that despite the free care policies for children under-5, families are sometimes obliged to pay for certain drugs such as antibiotics outside the PHCs, due to frequent drug shortages. Some authors have also noted other irregularities and effects in the implementation of these free healthcare policies (13,14), such as undeclared parallel expenditure or corrupt practices revealed by the Onwujekwe O. et al. research in Anglophone West Africa (37). The medium-term consequence is an increase in healthcare costs for families (36,38).

With regard to the limitations of this study, the health districts were deliberately chosen according to the area of intervention of the NGOs in the AIRE consortium; PHCs are therefore not entirely representative of all PHCs in each country. Consequently, it is necessary to interpret our results at the level of the districts concerned under the format of a pilot study.

Another weakness is that the assessment of healthcare facilities was not based on a standard approach such as SARA (39) as the aim was to focus solely on IMCI for logistical and financial reasons.

Finally, the survey was conducted just after the border closures due to the Covid-19 pandemic, resulting in the organisation of remote interviewer training and potential differences in the understanding data collection with a potential effect on data quality. It can be assume that these challenges were worsened by the Covid-19 pandemic that has occurred in 2020 (40) with the possible exception of access to oxygen and other health equipment to manage health emergencies, which has improved thanks to numerous initiatives such as ACT-A(41). Nevertheless, these limitations do not preclude the interpretation of the main results providing an overall picture of the health system capacity before the implementation of AIRE operational activities.

## CONCLUSION

This study highlights the unavailability of POs at the PHC level and a significant shortage of POs at hospital level at the start of the Covid-19 pandemic, which occurred before the AIRE project was implemented. It also reveals the lack of qualified human resources, difficulties in transferring patients to hospital and access to oxygen. There are also economic and social inequities in access to care for children under-5 which need to be taken into account (30). This baseline assessment has guided the upgrading of health infrastructures capacity (health medical equipment, medicines, oxygen supply, …). Our study provides useful baseline data for a better understanding of the context, and challenges to address for reducing mortality in children under-5 in West Africa.

## ACKNOWLEDGMENTS

We thank all the children and their families who participated in the study, as well as the healthcare staff at the participating hospitals and the sites involved. We thank the field project staff and the AIRE Research Study Group*. We thank the Ministries of Health of the participating countries for their support.

**Acknowledgements: Children, families, UNITAID and The AIRE Research Study Group: Country investigators**: Ouagadougou, Burkina Faso: S. Yugbaré Ouédraogo (PI), V. M. Sanon Zombré (CoPI), Conakry, Guinea: M. Sama Cherif (CoPI), I. S. Diallo (CoPI), D. F. Kaba, (PI). Bamako, Mali: A. A. Diakité (PI), A. Sidibé, (CoPI). Niamey, Niger: H. Abarry Souleymane (CoPI), F. Tidjani Issagana Dikouma (PI). **Research coordinators & data centers: Inserm U1295, Toulouse 3 University, France**: H. Agbeci (Int Health Economist), L. Catala (Research associate), D. L. Dahourou (Research associate), S. Desmonde (Research associate), E. Gres (PhD Student), G. B. Hedible (Int research project manager), V. Leroy (research coordinator), L. Peters Bokol (Int clinical research monitor), J. Tavarez (Research project assistant), Z. Zair (Statistician, Data scientist). **CEPED, IRD, Paris, France**: S. Louart (process manager), V. Ridde (process coordination). **Inserm U1137, Paris, France** : A. Cousien (Research associate). **Inserm U1219**, EMR271 IRD, **Bordeaux University, France** : R. Becquet (Research associate), V. Briand (Research associate), V. Journot (Research associate). **PACCI, CHU Treichville, Abidjan, Côte d’Ivoire** : S. Lenaud (Int data manager), C. N’Chot (Research associate), B. Seri (Supervisor IT), C. Yao (data manager supervisor). **Consortium NGOs partners: Alima-HQ (consortium lead), Dakar, Sénégal**: G. Anago (Int Monitoring Evaluation Accountability And Learning Officer), D. Badiane (Supply chain manager), M. Kinda (Director), D. Neboua (Medical officer), P. S. Dia (Supply chain manager), S. Shepherd (referent NGO), N. di Mauro (Operations support officer), G. Noël (Knowledge broker), K. Nyoka (Communication and advocacy officer), W. Taokreo (Finance manager), O. B. Coulidiati Lompo (Finance manager), M. Vignon (Project Manager). **Alima, Conakry, Guinea**: P. Aba (clinical supervisor), N. Diallo (clinical supervisor), M. Ngaradoum (Medical Team Leader), S. Léno (data collector), A. T. Sow (data collector), A. Baldé (data collector), A. Soumah (data collector), B. Baldé (data collector), F. Bah (data collector), K. C. Millimouno (data collector), M. Haba (data collector), M. Bah (data collector), M. Soumah (data collector), M. Guilavogui (data collector), M. N. Sylla (data collector), S. Diallo (data collector), S. F. Dounfangadouno (data collector), T. I. Bah (data collector), S. Sani (data collector), C. Gnongoue (Monitoring Evaluation Accountability And Learning Officer), S. Gaye (Monitoring Evaluation Accountability And Learning Officer), J. P. Y. Guilavogui (Clinical Research Assistant), A. O. Touré (Country health economist), J. S. Kolié (Country clinical research monitor), A. S. Savadogo (country project manager). **Alima, Bamako, Mali**: F. Sangala (Medical Team Leader), M. Traore (Clinical supervisor), T. Konare (Clinical supervisor), A. Coulibaly (Country health economist), A. Keita (data collector), D. Diarra (data collector), H. Traoré (data collector), I. Sangaré (data collector), I. Koné (data collector), M. Traoré (data collector), S. Diarra (data collector), V. Opoue (Monitoring Evaluation Accountability And Learning Officer), F. K. Keita (medical coordinator), M. Dougabka (Clinical research assistant then Monitoring Evaluation Accountability And Learning Officer), B. Dembélé (data collector then Clinical research assistant), M. S. Doumbia (country health economist), G. D. Kargougou (country clinical research monitor), S. Keita (country project manager). **Solthis-HQ, Paris**: S. Bouille (NGO referent), S. Calmettes (NGO referent), F. Lamontagne (NGO referent). **Solthis, Niamey**: K. H. Harouna (clinical supervisor), B. Moutari (clinical supervisor), I. Issaka (clinical supervisor), S. O. Assoumane (clinical supervisor), S. Dioiri (Medical Team Leader), M. Sidi (data collector), K. Sani Alio (Country supply chain officer), S. Amina (data collector), R. Agbokou (Clinical research assistant), M. G. Hamidou (Clinical Research Assistant), S. M. Sani (Country health economist), A. Mahamane, Aboubacar Abdou (data collector), B. Ousmane (data collector), I Kabirou (data collector), I. Mahaman (data collector), I Mamoudou (data collector), M. Baguido (data collector), R. Abdoul (data collector), A. Sahabi (data collector), F. Seini (data collector), Z. Hamani (data collector), L-Y B Meda (Country clinical research monitor), Mactar Niome (country project manager), X. Toviho (Monitoring Evaluation Accountability And Learning Officer), I. Sanouna (Monitoring Evaluation Accountability And Learning Officer), P. Kouam (program officer). **Terre des hommes-HQ, Lausanne**: S. Busière (NGO referent), F. Triclin (NGO referent). **Terre des hommes, BF**: A. Hema (country project manager), M. Bayala (IeDA IT), L. Tapsoba (Monitoring Evaluation Accountability And Learning Officer), J. B. Yaro (Clinical reearch assistant), S. Sougue (Clinical reearch assistant), R. Bakyono (Country health economist), A. G. Sawadogo (Country clinical research monitor), A. Soumah (data collector), Y. A. Lompo (data collector), B. Malgoubri (data collector), F. Douamba (data collector), G. Sore (data collector), L. Wangraoua (data collector), S. Yamponi (data collector), S. I. Bayala (data collector), S. Tiegna (data collector), S. Kam (data collector), S. Yoda (data collector), M. Karantao (data collector), D. F. Barry (Clinical supervisor), O. Sanou (clinical supervisor), N. Nacoulma (Medical Team Leader), N. Semde (clinical supervisor), I. Ouattara (Clinical supervisor), F. Wango (clinical supervisor), Z. Gneissien (clinical supervisor), H. Congo (clinical supervisor). **Terre des hommes, Mali**: Y. Diarra (clinical supervisor), B. Ouattara (clinical supervisor), A. Maiga (data collector), F. Diabate (data collector), O. Goita (data collector), S. Gana (data collector), S. Diallo (data collector), S. Sylla (data collector), D. Coulibaly (Tdh project manager), N. Sakho (NGO referent). **Country SHS team: Burkina Faso**: K. Kadio (consultant and research associate), J. Yougbaré (data collector), D. Zongo (data collector), S. Tougouma (data collector), A. Dicko (data collector), Z. Nanema (data collector), I. Balima (data collector), A. Ouedraogo (data collector), A. Ouattara (data collector), S. E. Coulibaly (data collector). **Guinea**: H. Baldé (consultant and research associate), L. Barry (data collector), E. Duparc Haba (data collector). **Mali**: A. Coulibaly (consultant and research associate), T. Sidibe (data collector), Y. Sangare (data collector), B. Traore (data collector), Y. Diarra (data collector). **Niger**: A. E. Dagobi (consultant and research associate), S. Salifou (data collector), B. Gana Moustapha Chétima (data collector), I. H. Abdou (data collector)

## Contributors

VL and VR conceptualised the research. The AIRE Research Study Group conducted training, data collection and management. HGB and AG with contributions from ZZ and VL realise the data analysis. HGB prepared the first draft of this article. All authors were involved in data interpretation and review of the final manuscript. VL is the guarantor to submit the manuscript.

## Data Availability Statement

The datasets generated and analysed during the current study are not publicly available. Access to processed deidentified participant data will be made available to any third Party after the publication of the main AIRE results stated in the Pan African Clinical Trial Registry Study statement (PACTR202206525204526, registered on 06/15/2022), upon a motivated request (concept sheet), and after the written consent of the AIRE research coordinator (Valeriane Leroy, Inserm U1295 Toulouse, France, orcid.org/0000-0003-3542-8616) obtained after the approval of the AIRE publication committee, if still active.

## REFERENCES

1. UN-IGME-Child-Mortality-Report-2018.pdf [Internet]. [cité 15 sept 2023]. Disponible sur: https://www.unicef.org/media/47626/file/UN-IGME-Child-Mortality-Report-2018.pdf

2. World Health Organization et UNICEF. Handbooklll: IMCI integrated management of childhood illness [Internet]. World Health Organization et UNICEF; 2005 [cité 13 sept 2023]. Disponible sur: https://apps.who.int/iris/handle/10665/42939

3. World Health Organization. Integrated Management of Childhood Illness: distance learning course [Internet]. Geneva: World Health Organization; 2014 [cité 10 sept 2023]. 15 p. Disponible sur: https://apps.who.int/iris/handle/10665/104772

4. World Health Organization. Revised WHO classification and treatment of pneumonia in children at health facilities: evidence summaries [Internet]. Geneva: World Health Organization; 2014 [cité 10 sept 2023]. 26 p. Disponible sur: https://apps.who.int/iris/handle/10665/137319

5. Wang H, Abajobir AA, Abate KH, Abbafati C, Abbas KM, Abd-Allah F, et al. Global, regional, and national under-5 mortality, adult mortality, age-specific mortality, and life expectancy, 1970–2016: a systematic analysis for the Global Burden of Disease Study 2016. The Lancet. 16 sept 2017;390(10100):10841Z150.

6. World Health Organization. Oxygen therapy for children: a manual for health workers [Internet]. Geneva: World Health Organization; 2016 [cité 24 août 2023]. 57 p. Disponible sur: https://apps.who.int/iris/handle/10665/204584

7. Gamponia MJ, Babaali H, Yugar F, Gilman RH. Reference values for pulse oximetry at high altitude. Archives of Disease in Childhood. 1 mai 1998;78(5):4611Z5.

8. Floyd J, Wu L, Hay Burgess D, Izadnegahdar R, Mukanga D, Ghani AC. Evaluating the impact of pulse oximetry on childhood pneumonia mortality in resource-poor settings. Nature. déc 2015;528(7580):S531Z9.

9. Subhi R, Adamson M, Campbell H, Weber M, Smith K, Duke T. The prevalence of hypoxaemia among ill children in developing countries: a systematic review. The Lancet Infectious Diseases. 1 avr 2009;9(4):2191Z27.

10. Graham H, Bakare AA, Ayede AI, Oyewole OB, Gray A, Peel D, et al. Hypoxaemia in hospitalised children and neonates: A prospective cohort study in Nigerian secondary-level hospitals. eClinicalMedicine. 1 nov 2019;16:511Z63.

11. Nair H, Nokes DJ, Gessner BD, Dherani M, Madhi SA, Singleton RJ, et al. Global burden of acute lower respiratory infections due to respiratory syncytial virus in young children: a systematic review and meta-analysis. Lancet. 1 mai 2010;375(9725):15451Z55.

12. Sarrassat S, Lewis JJ, Some AS, Somda S, Cousens S, Blanchet K. An Integrated eDiagnosis Approach (IeDA) versus standard IMCI for assessing and managing childhood illness in Burkina Faso: a stepped-wedge cluster randomised trial. BMC Health Serv Res. 16 avr 2021;21:354.

13. Ridde V, Diarra A, Moha M. User fees abolition policy in Niger: Comparing the under five years exemption implementation in two districts. Health Policy. 1 mars 2011;99(3):2191Z25.

14. Barroy H, Kutzin J, Coulibaly S, Bigeard A, Yaméogo SP, Caremel JF, et al. Public Financial Management as an Enabler for Health Financing Reform: Evidence from Free Health Care Policies Implemented in Burkina Faso, Burundi, and Niger. Health Systems & Reform. 1 janv 2022;8(1):e2064731.

15. Ridde V, de SARDAN JPO, Brouillet P, de PERETTI C. Abolishing user fees for patients in West Africa: lessons for public policy. mai 2013;

16. Johnson A, Goss A, Beckerman J, Castro A. Hidden costs: The direct and indirect impact of user fees on access to malaria treatment and primary care in Mali. Social Science & Medicine. 1 nov 2012;75(10):17861Z92.

17. Soucat A, Levy-Bruhl D, Gbedonou P, Drame K, Lamarque JP, Diallo S, et al. Local cost sharing in Bamako Initiative systems in Benin and Guinea: assuring the financial viability of primary health care. Int J Health Plann Manage. juin 1997;12 Suppl 1:S109–135.

18. Population des divisions administratives de la Guinée [Internet]. 2023 [cité 16 nov 2023]. Disponible sur: https://population.insguinee.org/resultat/

19. World Health Organization. Standards for improving quality of maternal and newborn care in health facilities [Internet]. Geneva: World Health Organization; 2016 [cité 26 mars 2024]. 73 p. Disponible sur: https://iris.who.int/handle/10665/249155

20. World Health Organization. Standards for improving the quality of care for children and young adolescents in health facilities [Internet]. Geneva: World Health Organization; 2018 [cité 26 mars 2024]. Disponible sur: https://iris.who.int/handle/10665/272346

21. Hedible GB, Louart S, Neboua D, Catala L, Anago G, Sawadogo AG, et al. Evaluation of the routine implementation of pulse oximeters into integrated management of childhood illness (IMCI) guidelines at primary health care level in West Africa: the AIRE mixed-methods research protocol. BMC Health Serv Res. 24 déc 2022;22:1579.

22. Alwadhi V, Bajpayee D, Kumar N, Mohanty JS, Mukherji K, Saboth PK, et al. E-IMNCI: a novel clinical diagnostic support system approach to strengthen effectiveness and quality of IMNCI implementation in India. BMJ Open Qual. 1 oct 2023;12(Suppl 3):e001857.

23. Graham HR, Olojede OE, Bakare AA, Iuliano A, Olatunde O, Isah A, et al. Measuring oxygen access: lessons from health facility assessments in Lagos, Nigeria. BMJ Glob Health. 3 août 2021;6(8):e006069.

24. McCollum ED, King C, Colbourn T, Graham H, Bernstein M, Wilson IH, et al. Pulse oximetry in paediatric primary care in low-income and middle-income countries. Lancet Respir Med. déc 2019;7(12):10011Z2.

25. World Health Organization, United Nations Children’s Fund (UNICEF). WHO-UNICEF technical specifications and guidance for oxygen therapy devices [Internet]. Geneva: World Health Organization; 2019 [cité 19 sept 2023]. (WHO medical device technical series;). Disponible sur: https://apps.who.int/iris/handle/10665/329874

26. King C, Boyd N, Walker I, Zadutsa B, Baqui AH, Ahmed S, et al. Opportunities and barriers in paediatric pulse oximetry for pneumonia in low-resource clinical settings: a qualitative evaluation from Malawi and Bangladesh. BMJ Open. 30 janv 2018;8(1):e019177.

27. Willcox ML, Peersman W, Daou P, Diakité C, Bajunirwe F, Mubangizi V, et al. Human resources for primary health care in sub-Saharan Africa: progress or stagnation? Human Resources for Health. 10 sept 2015;13(1):76.

28. Lancet T. GBD 2017: a fragile world. The Lancet. 10 nov 2018;392(10159):1683.

29. Dussault G, Codjia L, Zurn P, Ridde V. [Investing in human resources for health in French-speaking Africa: the contribution of the Muskoka project.]. Sante Publique. 3 mars 2018;S1(HS):91Z17.

30. Bado AR, Appunni SS. Decomposing Wealth-Based Inequalities in Under-Five Mortality in West Africa. Iran J Public Health. juill 2015;44(7):9201Z30.

31. Liste des médicaments essentiels de la Guinée. 2016.

32. Liste des médicaments essentiels au Niger. 2018.

33. Liste des médicaments essentiels du Mali. 2019.

34. Liste des médicaments essentiels du Burkina Faso. 2020;

35. Diarra A. Mise en œuvre locale de l’exemption des paiements des soins au Niger. Évaluation dans les districts sanitaires. Afrique contemporaine. 2012;243(3):771Z93.

36. Olivier de Sardan JP, Ridde V. L’exemption de paiement des soins au Burkina Faso, Mali et Niger. Les contradictions des politiques publiques. Afrique contemporaine. 2012;243(3):111Z32.

37. Onwujekwe O, Agwu P, Orjiakor C, McKee M, Hutchinson E, Mbachu C, et al. Corruption in Anglophone West Africa health systems: a systematic review of its different variants and the factors that sustain them. Health Policy Plan. sept 2019;34(7):5291Z43.

38. Nabyonga Orem J, Mugisha F, Kirunga C, Macq J, Criel B. Abolition of user fees: the Uganda paradox. Health Policy and Planning. 1 sept 2011;26(suppl_2):ii411Z51.

39. AbouZahr C, Weldedawit MA, Betizazu S, Bloem P, Bose K, Bucagu M, et al. Mesurer la disponibilité et la capacité opérationnelle des services (SARA) Un outil d’évaluation des établissements de santé (OMS).

40. Gilbert M, Pullano G, Pinotti F, Valdano E, Poletto C, Boëlle PY, et al. Preparedness and vulnerability of African countries against importations of COVID-19: a modelling study. The Lancet. 14 mars 2020;395(10227):8711Z7.

41. Global Oxygen Alliance Launched to Boost Access to Lifesaving Oxygen [Internet]. [cité 28 mars 2024]. Disponible sur: https://www.theglobalfund.org/en/news/2023/2023-05-24-global-oxygen-alliance-launched-to-boost-access-to-lifesaving-oxygen/

